# Short-term Outcomes of Trabeculectomy with Mitomycin-C in a Tertiary Eye Hospital in Nepal: A Prospective Descriptive Study

**DOI:** 10.1101/2025.01.19.25320810

**Authors:** Thinley, Indhira Paudyal, Pratibha Lama Joshi

## Abstract

**Objective:** This study evaluated the short-term outcomes of trabeculectomy with mitomycin-C in glaucoma patients at Tilganga Institute of Ophthalmology, Kathmandu, Nepal, between June 2021 and May 2022.

**Methods:** This was a prospective descriptive study, conducted on 56 eyes of 51 consecutive glaucoma patients who underwent trabeculectomy with mitomycin-C (0.02%). Follow-up assessments were carried out on postoperative day 1, week 1, month 1, month 3, and month 6. Statistical analysis was done using SPSS version 20 software.

**Results:** The study included 38 male (74.5%) and 13 female (25.5%) patients with an average age of 43.50 ± 15.45 years. A significant reduction in median intraocular pressure (IOP) from 24.33 mmHg preoperatively to 12.00 mmHg at 6 months postoperatively was observed (P < 0.01). The median number of anti-glaucoma medications decreased from 3.0 preoperatively to 0.0 at all postoperative visits. Complete surgical success was achieved in 85.7% of eyes. Common complications included shallow anterior chamber, hypotony, and IOP elevation.

**Conclusion:** Trabeculectomy with mitomycin-C demonstrated high surgical success rate with low complication rates in the short term, making it an effective and safe treatment option for glaucoma patients not medically controlled. It may benefit glaucoma patients facing challenges with long-term anti-glaucoma medication use and frequent follow-up visits, thus improving the management outcomes of this chronic condition.

## Introduction

Glaucoma is the leading cause of irreversible blindness worldwide.(1) (2) The global prevalence of glaucoma is 3.54% of which primary open-angle glaucoma (POAG) accounts for 3.05% and primary angle-closure glaucoma (PACG) 0.50%.(3) In Asia, the prevalence is 3.54%, with POAG and PACG accounting for 2.34% and 0.73% respectively. (4)In Nepal the prevalence was 1.9% of which 1.24% was primary open-angle glaucoma 0.39% was primary angle-closure glaucoma and 0.15% was secondary glaucoma.(5)

Raised intraocular pressure (IOP) apart from being the most important risk factor for glaucomatous optic nerve damage, is the only modifiable risk factor. As such, all the current medical and surgical treatment modalities are directed towards controlling IOP within a safe range.(6) (7)

While conservative treatments include topical anti-glaucoma medications and laser trabeculoplasty, advanced unresponsive cases often require surgical intervention such as trabeculectomy to control IOP and slow the disease progression. Studies have shown that lower IOP levels slow the progression of glaucomatous damage, and trabeculectomy is effective when other treatments fail.(1) (2) (8) (9)

Trabeculectomy, first described in 1968, has evolved into the standard surgical procedure for managing glaucoma with medically uncontrolled IOP.(10) This surgical technique creates an artificial drainage system for aqueous humor thus helping in reducing IOP. Various modifications including the adjunct use of anti-fibrotic agents like 5-Fluorouracil(5-FU) and Mitomycin-C(MMC) have improved its success rate.(7) (11)

Despite its benefits, there are some variations in outcomes of trabeculectomy according to age, glaucoma type, and ethnicity.(6)

For patients in developing countries like Nepal, where eye care centers are limited and economic challenges are significant, trabeculectomy offers an effective and lasting solution, reducing the need for costly medications and frequent follow-up visits.

This study aims to assess the outcomes of trabeculectomy with MMC 0.02% in the Nepalese population presenting to Tilganga Institute of Ophthalmology, to better understand its efficacy and potential as a reliable treatment option for specific glaucoma patients.

## Materials and Methods

### Inclusion Criteria

- All glaucoma patients above 18 years of age, who underwent trabeculectomy at our center during the study period.
- For the patients undergoing trabeculectomy in both eyes during the study period, both eyes were included in the study.

### Exclusion Criteria

- Patients with no light perception (NPL) vision.
- Patients who had undergone previous incisional ocular surgeries except for phacoemulsification.
- Patients with a follow-up period of less than 6 months.

### Interventional Details

Written informed consent was obtained from the patients before surgical intervention. Postoperative follow-ups were conducted on the first day, and then at one week, one month, three months, and six months.

The trabeculectomy procedure involved retrobulbar anesthesia with lidocaine, preparation and draping of the eye, creation of a fornix-based conjunctival flap, application of 0.02% mitomycin-C for three minutes, creation of a partial thickness scleral flap, sclerotomy, peripheral iridectomy, and suturing. Topical steroids, antibiotics, and cycloplegics were prescribed postoperatively.

### Outcome Measures

- **Primary Outcome:** Changes in intraocular pressure (IOP) was measured using a Goldman applanation tonometry. Preoperative baseline IOP was the average of IOP readings taken during the last three visits before surgery. Surgical success was defined as complete success-IOP ≤21 mmHg without anti-glaucoma medications (AGMs) or qualified success-IOP ≤21 mmHg with AGMs. Failure was defined as IOP >21 mmHg despite treatment.
- **Secondary Outcomes:** Number of anti-glaucoma medications, best corrected visual acuity (BCVA), and complications. BCVA was measured using Snellen’s visual acuity chart or Tumbling E chart later converted to log MAR units for data analysis.

### Data Collection

Data was collected using a standard pro forma, including patient details, demographic profiles, clinical findings, and management details. Data was sourced from patient charts, operation theatre (OT) notes, and electronic medical records (EMR) and subsequently stored in an electronic database secured with a password.

### Ethical Considerations

The study adhered to the principles of the Declaration of Helsinki. Informed consent was obtained from all the participants. Confidentiality was maintained by anonymizing patient data, and no patient was subjected to additional financial burdens due to participation in this study. The study protocol was approved by the Institutional Review Board of the National Academy of Medical Sciences (NAMS), Bir Hospital, Mahaboudha, Kathmandu, Nepal (Ref no. 136612078179).

### Data Analysis

Statistical analysis was performed using SPSS version 20. Non-parametric data was analyzed using the Wilcoxon Signed Rank Test. A p-value of less than 0.05 was considered significant. Results were expressed as percentages, mean ± standard deviation, and median for non-parametric variables, and presented in the form of tables, figures, and graphs.

## Results

A total of 56 eyes from 51 patients who underwent trabeculectomy with mitomycin-c (0.02 %) met the inclusion criteria. Thirty eight (74.5%) patients were male with the mean age of 43.50 ± 15.45 years. Regarding eye laterality, 33 (58.9%) eyes were left with 5 patients having both eyes included in the study. Fifty-one (91.1%) eyes were phakic and 5 (8.9%) were pseudophakic.

The median preoperative intraocular pressure was 24.33 mmHg (range: 12-66 mmHg). The median preoperative visual acuity in Log MAR units was 0.2 (range: 0.0-2.0). The median preoperative number of anti-glaucoma medications was 3.0 (range: 1-4).

The baseline demographic profile and clinical characteristics of the patients are summarized in **Table 1**.

**Table 1.** Demographic Profile and Clinical Characteristics.

| Characteristics | Value |
| --- | --- |
| No. of Patients | 51 |
| Male | 38 (74.5%) |
| Female | 13 (25.5%) |
| No. of Eyes | 56 |
| Right Eyes | 23 (41.1%) |
| Left Eyes | 33 (58.9%) |
| Lens Status |  |
| Phakic | 51 (91.1%) |
| Pseudophakic | 5 (8.9%) |
| Age (Mean ± SD) | 43.5 ± 15.5 years |
| Preoperative IOP (mmHg) | Median: 24.3 (12-66) |
| Preoperative Visual Acuity (LogMAR) | Median: 0.2 (0.0-2.0) |
| No. of Preoperative AGMs | Median: 3.0 (1-4) |

**Figure 1:**
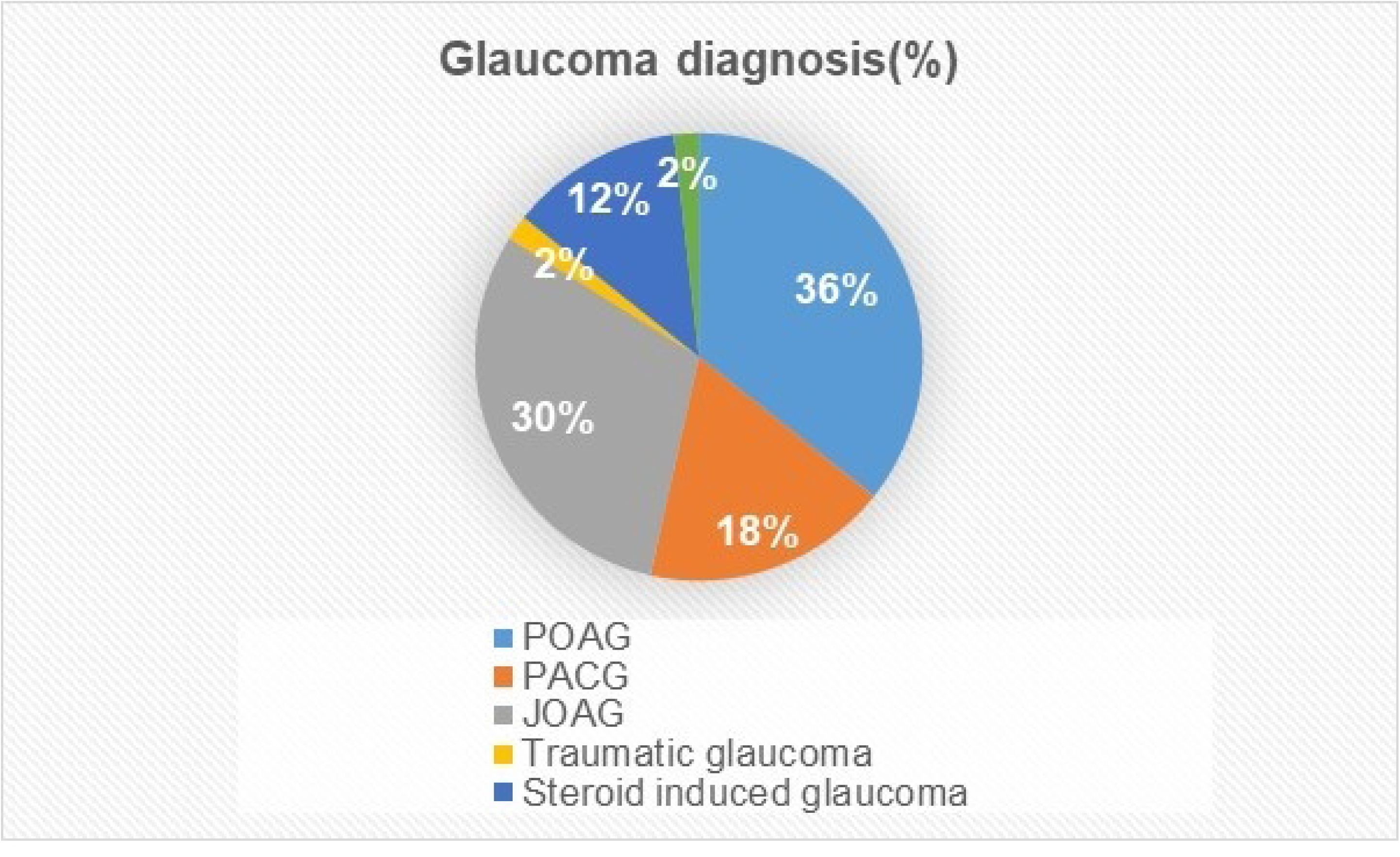
Distribution of eyes according to diagnosis. The majority (35.7%) of eyes were diagnosed with primary open-angle glaucoma (POAG), followed by juvenile open-angle glaucoma (JOAG) (30.4%). Primary angle-closure glaucoma (PACG) accounted for 17.9%, steroid-induced glaucoma for 12.5%, and traumatic and uveitic glaucoma each contributed 2%.

**Figure 2:**
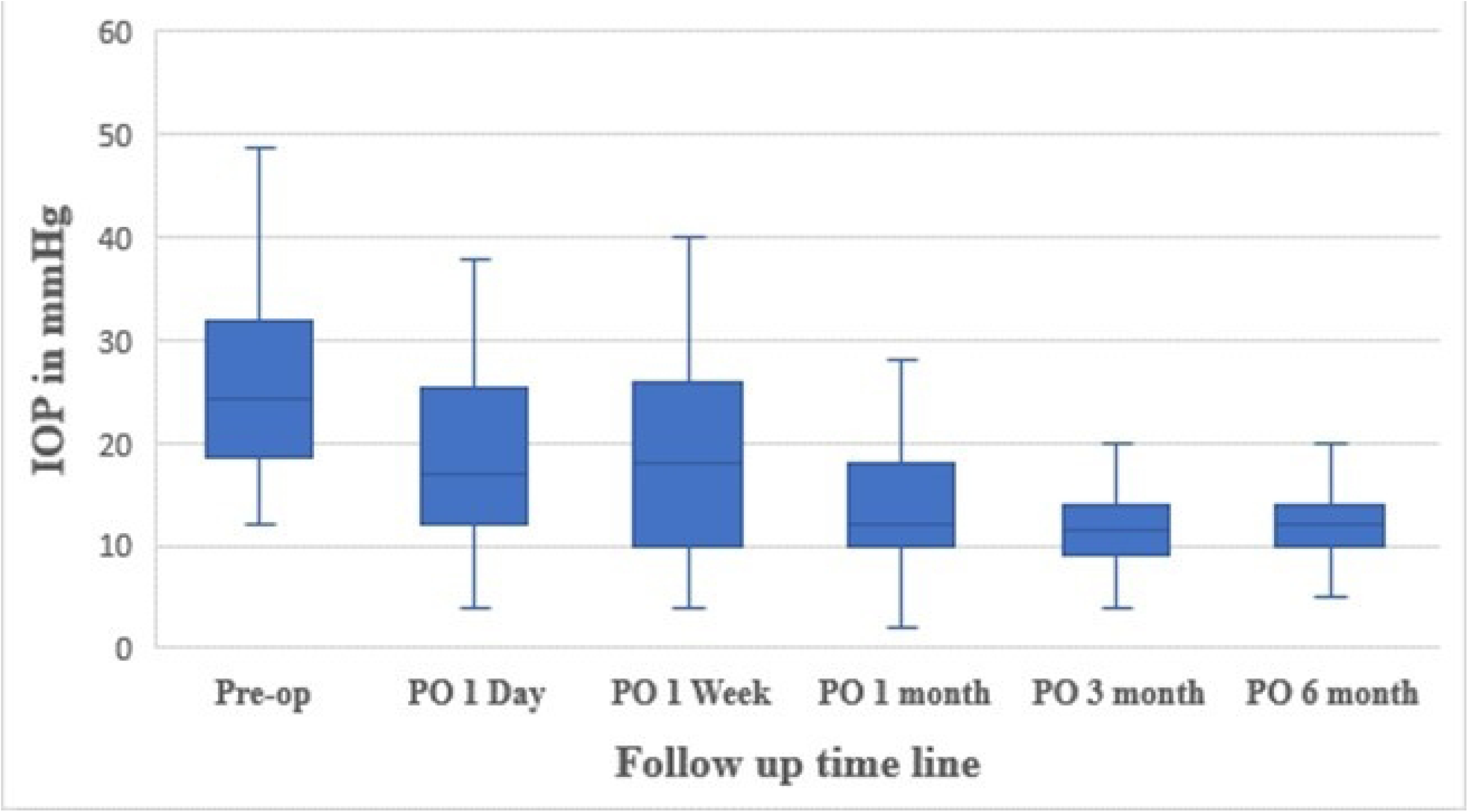
Pre- and post-operative IOP at various follow-up visits. Graphical representation of the comparison between preoperative median IOP and postoperative values at different time points. A significant and sustained reduction in median IOP is observed at all postoperative time points.

In the current study, at the last follow-up visit 85.7 % of the eyes that underwent trabeculectomy were free from anti-glaucoma medications (**Table 2**). The median number of anti-glaucoma medications used preoperatively was 3.0. At 6 months following surgery, five (8.9%) eyes required one anti-glaucoma drop, one (1.8%) eye required 2 anti-glaucoma drops, and two (3.6%) eyes required 3 drops to control the IOP within the target limits.

**Table 2.** Anti-glaucoma medications used at six months post-operatively.

| No. of medications | No. of eyes | Percent (%) |
| --- | --- | --- |
| 0 | 48 | 85.7 |
| 1 | 5 | 8.9 |
| 2 | 1 | 1.8 |
| 3 | 2 | 3.6 |

**Figure 3:**
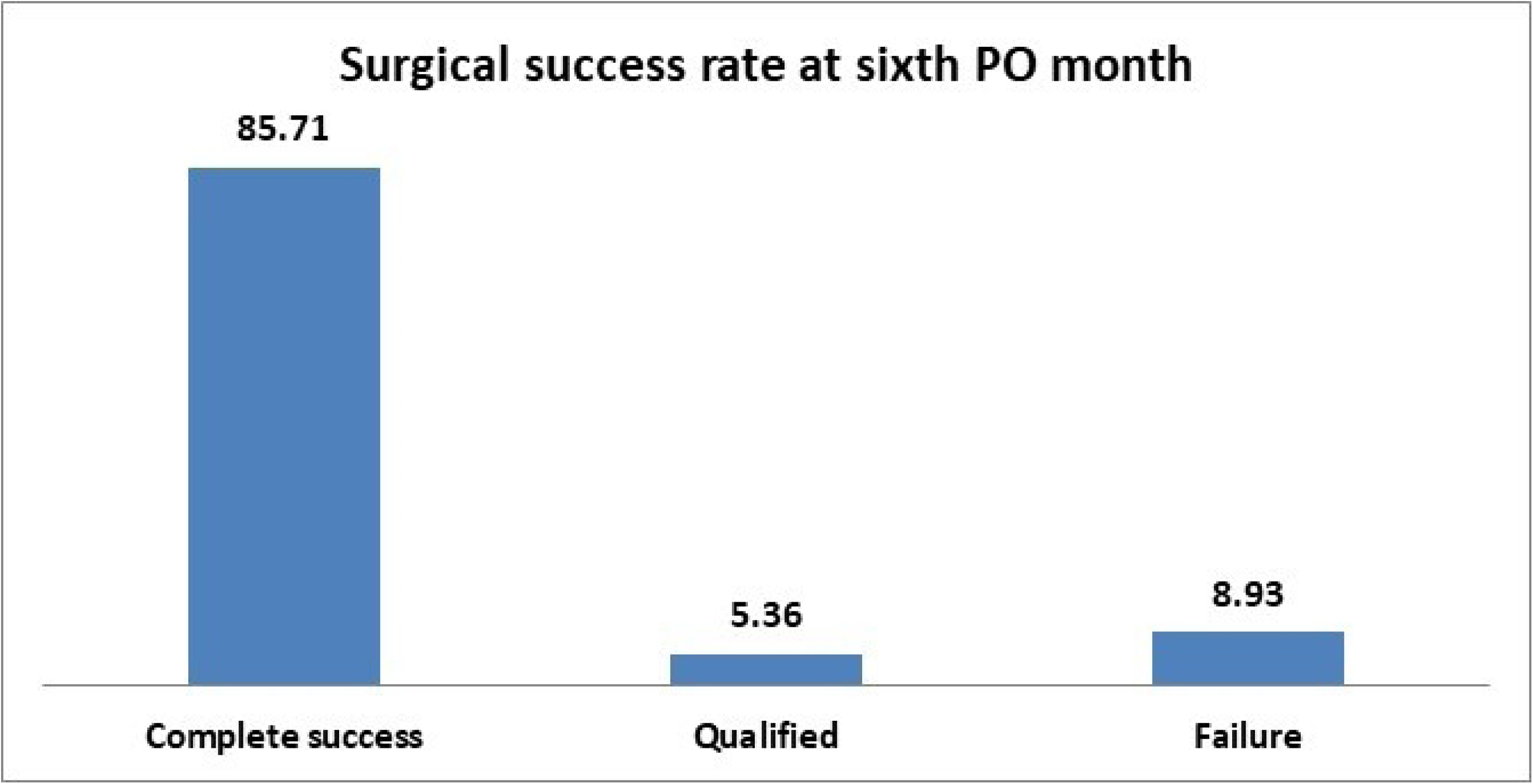
Bar chart showing the surgical success rates in percentage. At the final follow-up visit, complete surgical success was achieved in 48(85.71%) eyes, qualified success in 3 (5.36) eyes, and surgical failure in 5(8.93%) eyes. Three eyes required anti-glaucoma drops to maintain IOP below 21 mmHg. None of the eyes in the failure category required any additional interventions to control the IOP.

**Figure 4:**
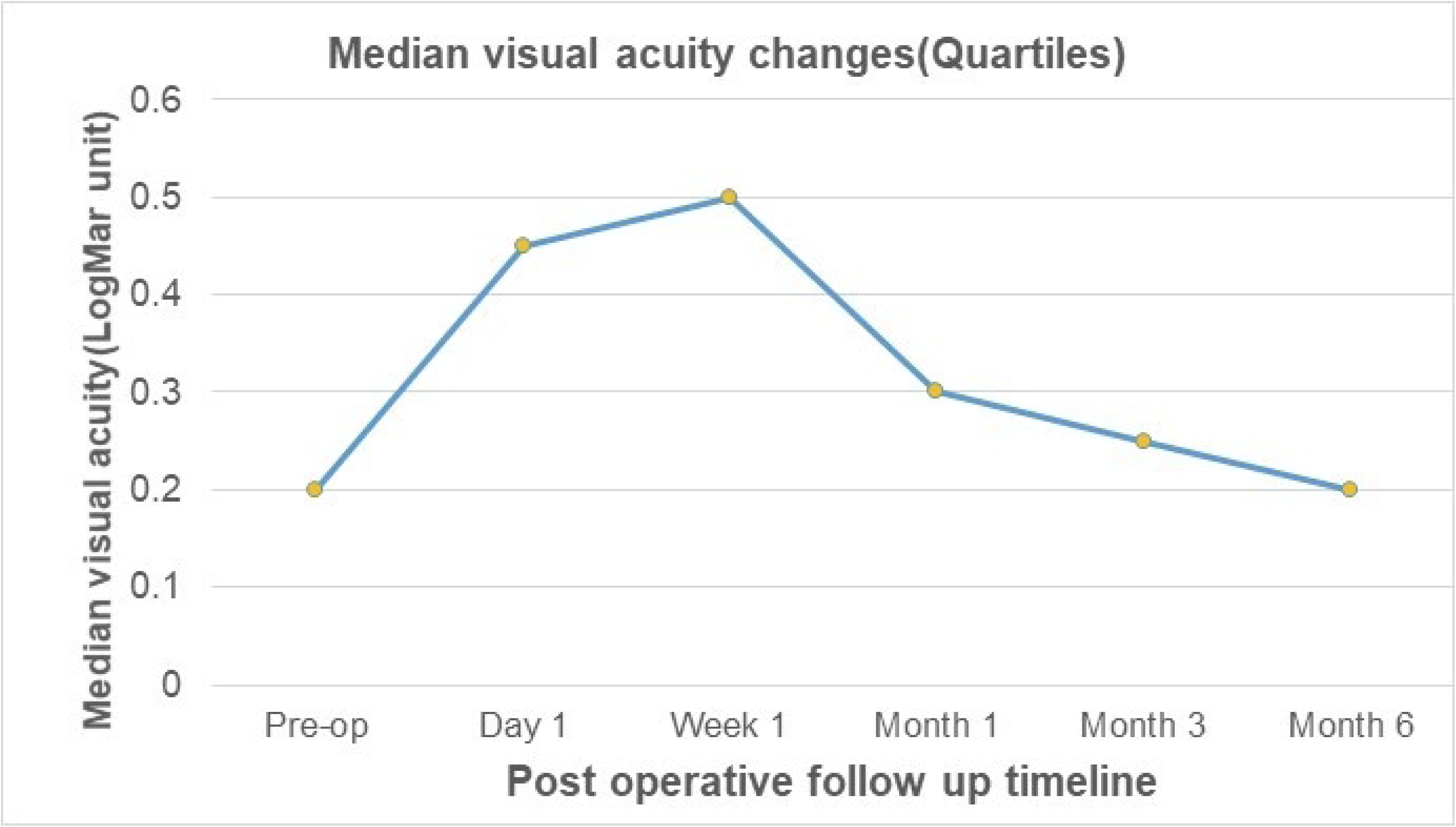
Postoperative visual acuity changes against pre-operative value. The median (quartiles) preoperative visual acuity (Log MAR) was 0.20(0.0, 0.3) and postoperative values were 0.45 (0.22, 0.80), 0.50 (0.30, 0.80), 0.30 (0.20, 0.50), 0.25 (0.0, 0.45) and 0.20 (0.0, 0.50) at day 1, week 1, month 1, month 3 and month 6 respectively. Visual acuity declined slightly in the early postoperative period but improved gradually, approaching the preoperative state by the last visit (p =0.045).

In this study, the more common postoperative complications were shallow anterior chamber, hypotony, and IOP elevation (**Table 3**). Shallow AC was present in 16.2%, hypotony in 39.2% and IOP elevation in 42.9%. Other less frequent complications included hypotony maculopathy(3.6%), bleb leak(7.1%), and choroidal detachment (3.6%). Cataract formation or progression was observed in five eyes (8.9%) during the follow-up, of which one eye had undergone cataract surgery during the study period. Iris obstructing the ostium was noted in 3(5.4%) eyes and all of them underwent iris repositioning and wound repair surgery. Blebitis and encapsulated bleb were noted in 1(1.8%) eye each during the study period.

**Table 3.**
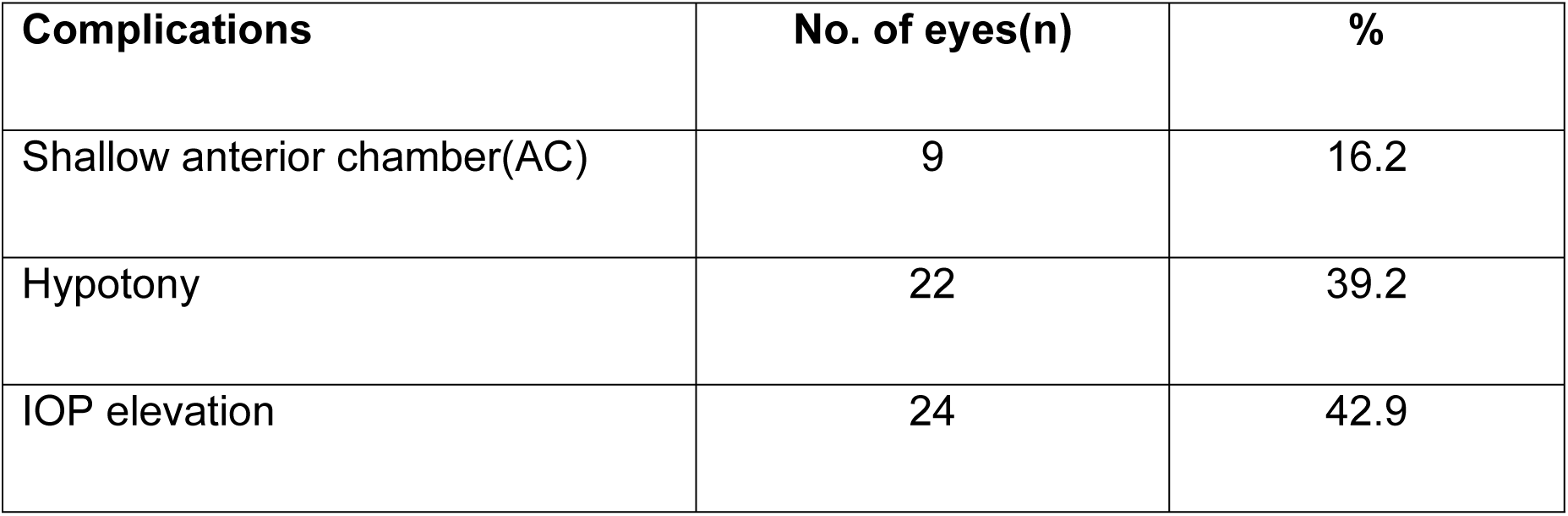
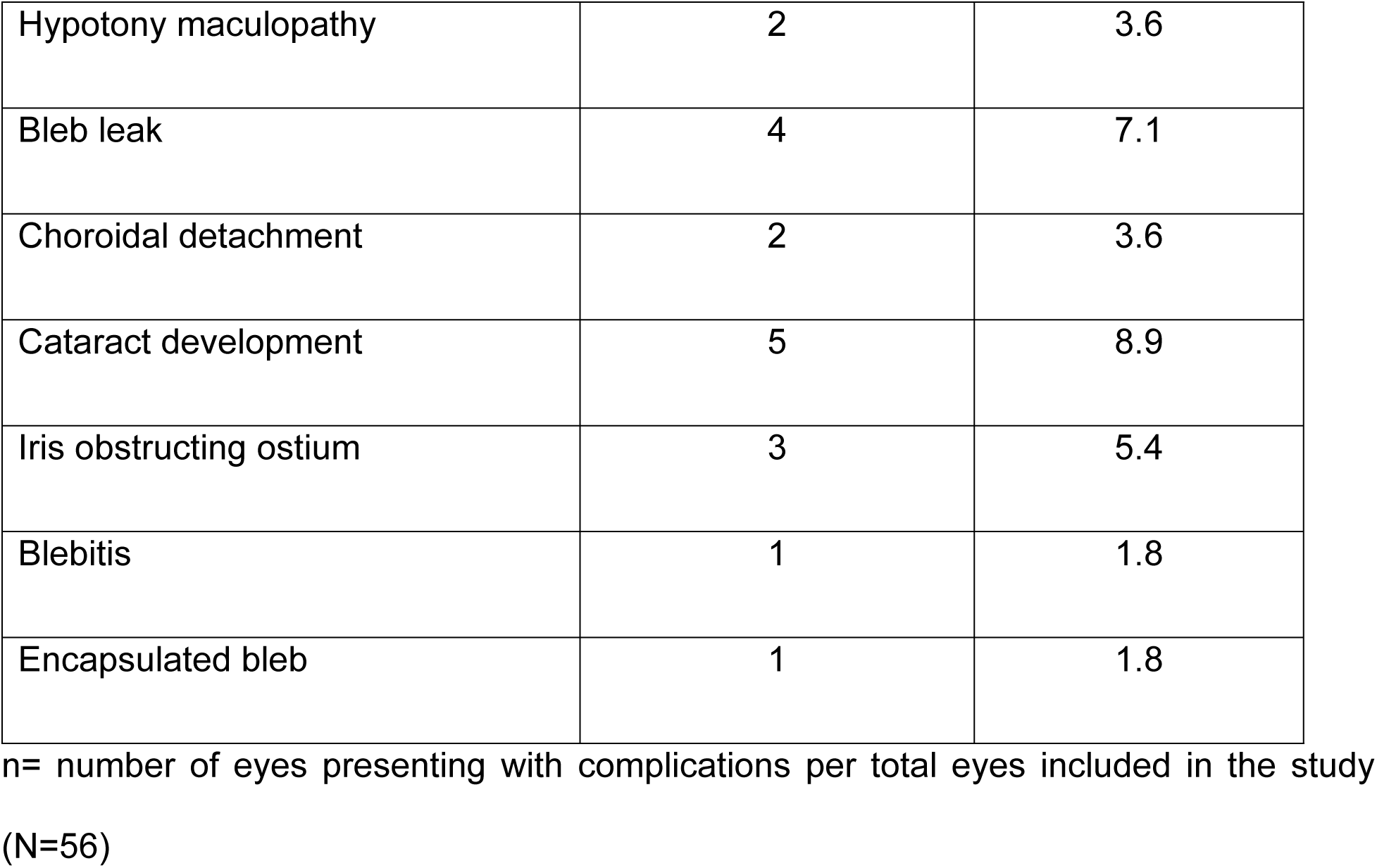
Post-operative complications at different post-operative visits.

| Complications | No. of eyes(n) | % |
| --- | --- | --- |
| Shallow anterior chamber(AC) | 9 | 16.2 |
| Hypotony | 22 | 39.2 |
| IOP elevation | 24 | 42.9 |
| Hypotony maculopathy | 2 | 3.6 |
| Bleb leak | 4 | 7.1 |
| Choroidal detachment | 2 | 3.6 |
| Cataract development | 5 | 8.9 |
| Iris obstructing ostium | 3 | 5.4 |
| Blebitis | 1 | 1.8 |
| Encapsulated bleb | 1 | 1.8 |

n= number of eyes presenting with complications per total eyes included in the study (N=56)

## Discussion

Trabeculectomy is the most commonly performed operation for the surgical treatment of glaucoma. Numerous studies have demonstrated the safety and efficacy of this procedure for both short- and long-term control of IOP. (1) (2) (12)

The present study has demonstrated a significant decline in IOP following trabeculectomy surgery performed in uncontrolled glaucoma patients. In this study, the median intraocular pressure decreased significantly from 24.33(18.41, 32.00) mmHg preoperatively to 12.00(10.0, 14.0) mmHg at 6 months postoperatively (P value < 0.01). Similar results have been noted in other short-term studies(6) (13) (14) (15) (16) and similar efficacy in IOP reduction has been noted in other longer-duration studies.(12) (17) (18)

According to our results, the surgical success rate of trabeculectomy with MMC at six months postoperatively was 85.7 % (complete success) which corroborates with results of other studies.(6) (8) (19) (20) (21)

Similarly, the median (Quartiles) number of anti-glaucoma medications have decreased from 3.0(3.0, 4.0) preoperatively to 0.0(0.0, 0.0) at six months postoperatively. The majority of the patients (85.7%) were free from anti-glaucoma drops at the end of six months’ follow up period. This finding of our study correlates with the results of study by Scott I.et al(22) in which 88.2 % of patients were off medications at 1 year and 83.9 % at 2 years, although the follow up duration was relatively longer in their study. Likewise, other studies have also shown significant reduction in anti-glaucoma medications following trabeculectomy.(2) (16) (17)

Regarding the postoperative visual acuity, in our study the median value dropped from the preoperative value during the early post-operative period, however, the visual acuity returned to near preoperative value at the end of sixth months’ follow-up period (P<0.045). These results are similar to what was observed in a study conducted by Stead R.(12) and King and Sirisawad P.(23). Some other studies have also reported similar results.(13)(15) (24)

In the present study, the more commonly observed postoperative complications were shallow anterior chamber, hypotony, and IOP elevation. Our results correlate with the findings of the study by Abe R.et al(25) and Jampel et al.(26)

Other studies(1) (16) (17) (27) have reported hyphema as a common complication, surprisingly no case of hyphema was recorded in the present study.

IOP elevation was another important complication noted in 24(42.9 %) eyes in our study at various postoperative time points. Daher F. et al (28) have reported IOP spikes as complication in their study, however their observation was noted within the first postoperative hours following trabeculectomy.

Other complications including hypotony maculopathy, bleb leak, choroidal detachment, cataract formation or progression, and iris obstructing the ostium, were noted with less frequency in our study. Almobarak F. et al.(29) have reported a few cases of a delayed bleb-related endophthalmitis, but in our study, none of the eyes had endophthalmitis as complication during the study period.

## Limitations

There are several limitations to this study. Firstly, the follow-up period was limited to six months, which may not capture long-term outcomes and complications. Secondly, the study was conducted at a single institution and the sample size is also relatively small, which may limit the generalizability of the results to other settings or populations. Additionally, the study design was descriptive and did not include a control group for comparison. Finally, self-reported adherence to postoperative medication and follow-up visits could have introduced some bias.

## Recommendations

Despite these limitations, the study provides valuable insights into the short-term efficacy and safety of trabeculectomy with mitomycin-C in a Nepalese population. Future research with longer follow-up periods, multicenter designs, and control groups is needed to confirm these findings and explore their applicability in different patient populations and contexts.

## Conclusion

Trabeculectomy with mitomycin-C is an effective surgical procedure for lowering IOP in uncontrolled glaucoma patients, with minimal complications and a good safety profile. This study reinforces the procedure’s role in glaucoma management, especially in developing countries where access to medical care and resources is limited.

## Data Availability

All relevant data are within the manuscript and its Supporting Information files.

## Acknowledgements

We would like to extend our gratitude to Mr. Suman S. Thapa, glaucoma specialist, who used to work at Department of Galucoma at Tilganga Institute of Ophthalmology then for his value insights and directions for this study at its initial stage.

